# Epidemic Model Guided Machine Learning for COVID-19 Forecasts in the United States

**DOI:** 10.1101/2020.05.24.20111989

**Authors:** Difan Zou, Lingxiao Wang, Pan Xu, Jinghui Chen, Weitong Zhang, Quanquan Gu

## Abstract

We propose a new epidemic model (SuEIR) for forecasting the spread of COVID-19, including numbers of confirmed and fatality cases at national and state levels in the United States. Specifically, the SuEIR model is a variant of the SEIR model by taking into account the untested/unreported cases of COVID-19, and trained by machine learning algorithms based on the reported historical data. Besides providing basic projections for confirmed and fatality cases, the proposed SuEIR model is also able to predict the peak date of active cases, and estimate the basic reproduction number (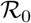). In particular, the forecasts based on our model suggest that the peak date of the US, New York state, and California state are 06/01/2020, 05/10/2020, and 07/01/2020 respectively. In addition, the estimated 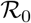 of the US, New York state, and California state are 2.5, 3.6 and 2.2 respectively. The prediction results for all states in the US can be found on our project website: https://covid19.uclaml.org, which are updated on a weekly basis, and have been adopted by the Centers for Disease Control and Prevention (CDC) for COVID-19 death forecasts (https://www.cdc.gov/coronavirus/2019-ncov/covid-data/forecasting-us.html).

## 1 Introduction

The novel coronavirus disease (COVID-19), an infectious disease caused by severe acute respiratory syndrome coronavirus 2 (SARS-CoV-2) (Chan et al., 2020; WHO, 2020b), has emerged into a global pandemic and lead to 233,839 death toll in the world as of April 30, 2020 (WHO, 2020a). The treatments for COVID-19 are still under investigation and in early stages. There are no specific vaccines or medicines that showed significant effectiveness on COVID-19 so far. As a consequence, one of the best ways to prevent the spread of COVID-19 in the short term is to follow the mitigation strategies such as social distancing, quarantine, and isolation. For example, the state of California in the US has issued mandatory stay-at-home order on March 19, shutting down all non-essential businesses. Only essential services, such as grocery stores, pharmacies, delivery restaurants, have remained open, and residents who need to leave home to take part in essential activities are advised to practice social distancing.

With the increasing availability of public data on COVID-19, more and more researches (Flaxman et al., 2020; Bendavid et al., 2020; Sutton et al., 2020; Altieri et al., 2020; Bertozzi et al., 2020; Murray et al., 2020) have been carried out to understand and prevent the spread of COVID-19 from different aspects. Among them, one important research direction is to model and forecast the spread of COVID-19, such as predicting the peak of the active cases on the virus and the size of the coronavirus outbreak. Such results can help government agencies better understand the overall impact of the disease and also facilitate policy makers in terms of pandemic preparedness and response such as allocating the medical resources.

One widely used method for modeling the spread of infectious disease is to use epidemic models such as Susceptible-Infected-Resistant (SIR) (Kermack and McKendrick, 1927) and Susceptible-Exposed-Infected-Removed (SEIR) (Hethcote, 2000). Such epidemic models are quite useful in describing the dynamics of transmission and are well-suited for predicting the peak of active cases on the virus. From the decision-making perspective, the peak prediction is able to forewarn the health system when to expect a surge in cases. Furthermore, the reproduction number (Fraser et al., 2009) estimated by the epidemic model can be directly used to measure the effectiveness of the intervention strategies such as social distancing and quarantine.

Several recent work used epidemic models such as the SIR and SEIR models (Imai et al., 2020; Li et al., 2020a; Wu et al., 2020; Kucharski et al., 2020; Read et al., 2020; Tang et al., 2020; Ferguson et al., 2020) to simulate the spread of COVID-19 in different regions and were able to forecast the size and severity of such epidemic outbreak. Some other work (Chinazzi et al., 2020; Kraemer et al., 2020; Dandekar and Barbastathis, 2020) also applied these epidemic models to study the role of quarantine controls such as travel restrictions in the spread of COVID-19. Most of the aforementioned studies consider classical epidemic models, e.g., the SIR and SEIR models, and base their analyses on the public reported data. However, it is often the case that the number of publicly reported cases (including confirmed cases and recovered cases) are much less than their real numbers as many infectious cases have not been tested due to test capability and asymptomatic patients, or even possibly under-reporting (Li et al., 2020b). As a result, classical epidemic models such as SIR and SEIR cannot accurately characterize the epidemic evolution of COVID-19 without taking such unreported cases into consideration. In addition, most existing work is focused on the nation-wide prediction. Nevertheless, it is also very important and beneficial to provide state and county level forecasts to assist local public health departments and governments to prevent the spread of COVID-19.

The goal of this paper is to make good use of the current public data on COVID-19 to better understand the spread of the coronavirus and to facilitate informed decisions by policy makers. In order to achieve this goal, we develop a new epidemic model, called the SuEIR model, to forecast the active cases and deaths of COVID-19 by taking the untested/unreported cases into consideration. In addition, we use machine learning based methods to train our model, which enables us to train the model efficiently. Based on the proposed model, we are able to make accurate predictions on the numbers of confirmed cases and fatality cases for nation, states and and counties. Moreover, our model can also predict the peak dates of active cases and estimate the basic reproduction number (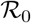) of different states in the US.

### 1.1 The SIR and SEIR Models

In this subsection, we briefly introduce two classical epidemic models, i.e., the SIR (Kermack and McKendrick, 1927) and SEIR models (Hethcote, 2000), which have been adopted in many previous work to study the epidemic outbreaks such as SARS (Fang et al., 2006; Saito et al., 2013; Smirnova et al., 2019) and the ongoing COVID-19 (Read et al., 2020; Tang et al., 2020; Wu et al., 2020).

The SIR model. The SIR model is an epidemic model that shows the change of infection rate over time. More specifically, it characterizes the dynamic interplay among the susceptible individuals (S), infectious individuals (I) and removed individuals (R) (including recovered and deceased) in a certain place. In the SIR model, the susceptible individuals may become infectious individuals over time, which depends on the spread rate of the virus, often called the contact rate. Recovered individuals are assumed to be immune to the virus and thus cannot become susceptible again. To characterize this dynamics, we use *S_t_, I_t_, R_t_* to represent the number of susceptible, infectious, and removed individuals at time *t*, respectively. Suppose that the total population in a certain area is fixed as *N*, then the evolving equations of the above variables over time are defined as follows:

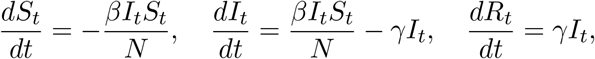

where *β* is the contact rate between the susceptible and infectious groups, and *γ* is the transition rate between the infectious and removed groups. The above ordinary differential equations indicate that at every time unit the total number of susceptible individuals will decrease by a quantity −*βI_t_S_t_/N*, which will transit into the infectious group. Apart from the increase from the transition of susceptible individuals, the size of the infectious group will also decrease by a factor of *γ*.

The SEIR model. For many diseases, there is often an incubation period during which individuals who have been exposed to the virus may not be as contagious as the infectious individuals. Therefore, it is necessary to separately model these cases as the “Exposed” group, and this gives rise to the SEIR model. The dynamics of the SEIR model introduces a new compartment *E_t_*, which models the number of individuals that are exposed to the disease but have not developed obvious symptoms. Among all the exposed cases, there are only a *σ* fraction of people who will develop observable symptoms in a time unit. Therefore, the dynamic of this model can be defined by the following ordinary differential equations:

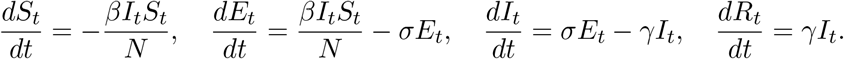

Compared with the SIR model, the SEIR model has more elaborated model parameters. The parameters *σ, β* and *γ* can be learned from the reported data.

Reproduction number. An important quantity to characterize the dynamic of a pandemic is the basic reproduction number 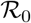, which is the expected number of cases directly generated by one case in a population where all individuals are susceptible to infection (Fraser et al., 2009). The basic reproduction number in the SIR and SEIR models can be computed as 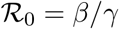.

## 2 Methods

In this section, we propose a new epidemic model and a machine learning method to train this model.

### 2.1 The SuEIR Model

It is observed that COVID-19 has an incubation period ranging from 2 to 14 days (Lauer et al., 2020). It has also been observed that individuals who have been exposed to the coronavirus can also infect the susceptible group during this period. In addition, it is often the case that the number of reported cases (including confirmed cases and recovered cases) are less than their real numbers as many exposed cases have not been tested, which will not pass to the next compartment. However, such important factors cannot be characterized by the classical epidemic models such as the SIR and SEIR models. We also observe that directly applying SIR or SEIR model to fit the reported data will lead to unreasonable predictions. Therefore, we proposed a new epidemic model that takes the untested/unreported cases as well as the “silent spreaders” into consideration. We call our model the SuEIR model and it is illustrated in Figure 1.

**Figure 1:**
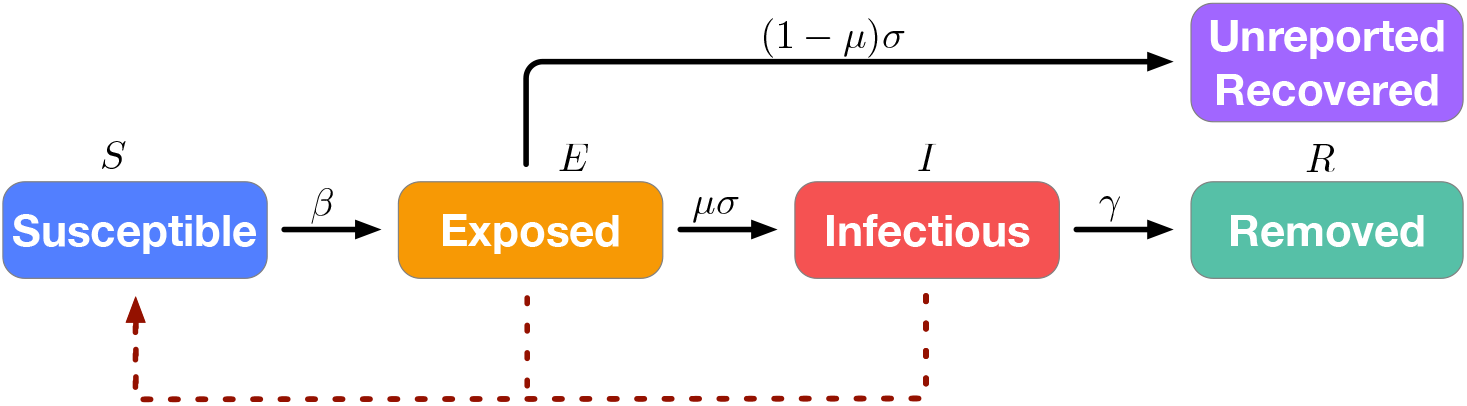
Illustration of the SuEIR model. Solid lines represent the transitions of individuals and dashed lines represent the routes of infection.

In particular, the compartment Exposed in our model is considered as the individuals that have already been infected and have not been tested. Therefore, they also have the capability to infect the susceptible individuals. Moreover, some of such individuals can receive a test and be further passed to the Infectious compartment (as well as reported to the public), while the others will recover but not appear in the publicly reported cases. Therefore, we introduce a new parameter 0 *< μ <* 1 in the evolution dynamics of *I_t_* to characterize the ratio of the exposed cases that are confirmed and reported to the public, which we call it the *discovery rate*. This discovery rate reflects the unreported/undiscovered cases, which is an important latent factor in the dynamics of the epidemic model. As a result, we propose to use the following ordinary differential equations to describe our proposed SuEIR model:

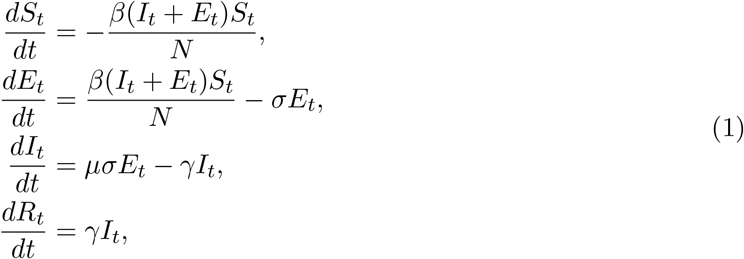

where *β* denotes the contact rate between the susceptible and “infected” groups (including both exposed and infectious compartments in Figure 1), *σ* denotes the ratio of cases in the exposed compartments that are either confirmed as infectious or dead/recovered without confirmation, *μ* is the discovery rate of the infected cases, and *γ* denotes the transition rate between the compartments *I* and *R*.

### 2.2 Parameter Learning for the SuEIR Model

In this subsection, we will introduce our proposed machine learning method for training the SuEIR model. In addition, we will also present the detailed configurations used in our experiments.

Model training. As aforementioned in this paper, our model can be described by the ODE (1), which is determined by the parameters ***θ*** = (*β*, *σ*, *γ*, *μ*). In particular, given the model parameters ***θ*** and initial quantities *S*_0_, *E*_0_, *I*_0_, and *R*_0_, we can compute the number of individuals in each group (i.e., *S*, *E*, *I*, and *R*) at time *t*, denoted by 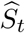, 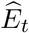, 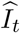 and 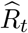, via applying standard numerical ODE solvers onto the ODE (1). Then we propose to learn the model parameter 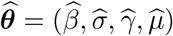 by minimizing the following logarithmic-type mean square error (MSE):

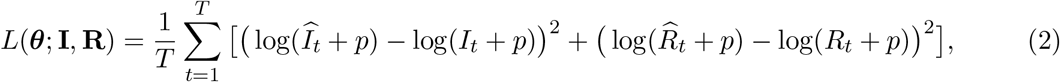

where 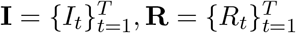 with *I_t_* and *R_t_* denote the reported numbers of infected cases and removed cases (including both recovered cases and fatality cases) at time t (i.e., date), and p is the smoothing parameter used to ensure numerical stability. Note that given *S*_0_, *E*_0_, *I*_0_ and *R*_0_, 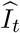 and 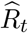 can be described as differentiable functions of the parameter ***θ***. Then the model parameter 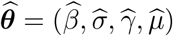 can be learnt by applying standard gradient based optimizer (e.g., BFGS) onto the loss function (2) under the constraint that *β, σ, γ, μ ∊[0,1]*.

Estimation of the number of removed cases *R_t_*.

Note that *I_t_* and *R_t_* in our model determine the number of “current” infectious cases (a.k.a., active cases) and the removed cases, i.e., the sum of recovered and fatality cases, respectively. However, most of the reported data only include the number of confirmed cases, i.e., sum of infected cases and removed cases *I_t_* + *R_t_*. In order to train the model, we need to get *I_t_* and *R_t_* separately. In addition, the SuEIR model can only predict the number of removed cases, while in many cases people are more interested in the number of fatality cases. Therefore, in order to enable the training of the SuEIR model, as well as provide the predictions for the number of fatality cases, we have to: (1) estimate the number of removed cases; (2) determine the number of active cases in the reported data by subtracting the estimated number of removed cases. In order to do so, we propose to use the following exponential function to model the ratio between the daily increased fatality cases and the removed cases,

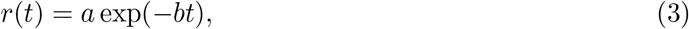

where *a,b >* 0 are parameters controlling the shape of the exponential function and *t* denotes the number of days since the starting date. In order to demonstrate its effectiveness, we evaluate the approximation error based on the reported data in four countries: US, China, France, and Italy, which have separately reported fatality and recovered cases. More specifically, given parameters *a, b* and the number of fatality cases, we are able to estimate the corresponding number of removed cases. Then the optimal parameters *a* and *b* are obtained by minimizing the MSE between the reported number of removed cases and the estimated one on different dates. The results are displayed in Figure 2, which clearly shows that the exponential functions can well describe the ratio between the daily increased numbers of fatality and removed cases. For each state and county in the US, we try several different choices of *a* and *b* around the optimal ones we obtained for the US, and pick the one with the smallest validation error.

**Figure 2:**
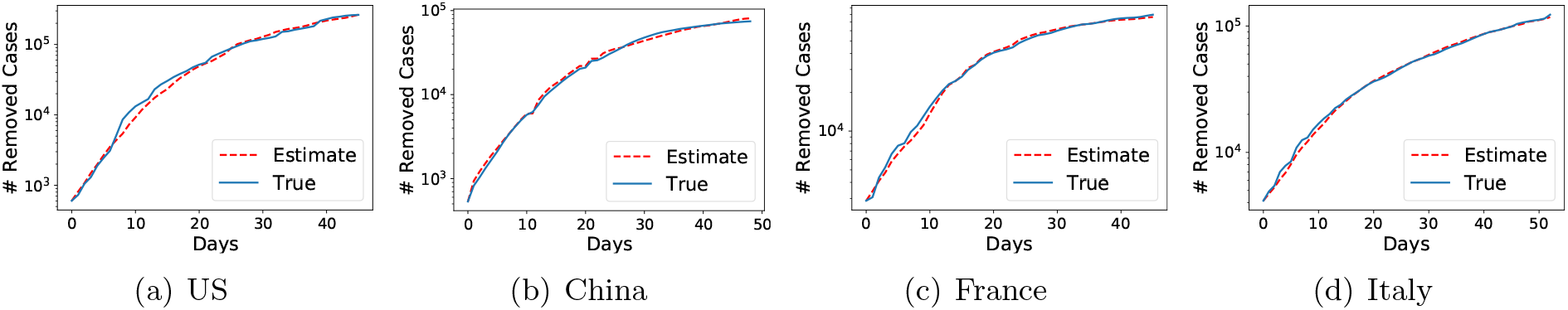
Estimated number of removed cases for different countries.

Initialization. In terms of the initialization, we directly set 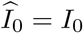 and 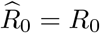^1^. Additionally, one can typically set 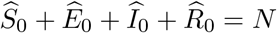, where *N* is the total population of the region (which can be either a country or a state/county). However, since most of the states/counties in the US have already issued the stay-at-home offer, the actual total number of cases in the SuEIR model will be strictly less than *N*. Thus we set 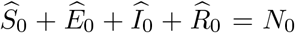 for some *N_0_ < N*. Moreover, it is worth noting that the initialization of *E*, i.e., 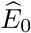, is a bit tricky since we do not know the number of infected cases before testing them. It is not reasonable to set 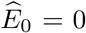 since generally there has already existed a large number of infected cases when the local governments began to test. Therefore, we propose to use a validation set to choose the optimal initial estimates *N*_0_ and 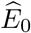 when training our model.

Validation set. To determine the initial values of *N*_0_ and *E*_0_, we first divide our data into the training data set and the validation data set. In detail, we choose the data in the most recent 7 days as the validation set, while treating the remaining as the training set. For example, suppose we have the data up to May 10, 2020, the data after May 3, 2020 will be used as the validation set, and the data up to May 3, 2020 will be used as the training set. We then do a grid search on different combinations of *N*_0_ and *E*_0_ and train different models on the training set accordingly. Finally, we choose the combination of N_0_ and E_0_ with the smallest validation loss (evaluated using the loss function (2)) along with the best model parameters (i.e., P, 7, a, *y*) to build the SuEIR model for prediction.

### 2.3 Confidence Interval

Given the initial quantities *S_0_, E_0_, I_0_, R_0_*, we can solve the optimization problem in (2) to obtain the model parameter 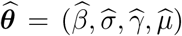. To assess the confidence of our estimator, we construct the confidence interval of ***θ*** following the previous work (Ma, 2020). More specifically, for a valid model parameter ***θ***, we can compute the loss *L*(***θ***) in (2), and construct the test statistic as 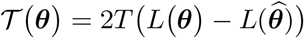, which represents the loglikelihood ratio between the point estimator 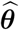 and ***θ***. Note that ***θ*** contains four free parameters (i.e., *β*, *σ, γ* and *μ*) while 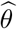 is fixed. By Wilks’s Theorem (Wilks, 1938), we know that 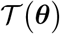 follows 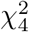 distribution asymptotically. As a result, we can compare 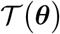 with the (1 − *α*) quantile of the 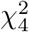 distribution and determine whether ***θ*** is in the confidence interval or not. In our experiment, we apply grid search on both sides of the point estimator 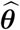 to find the boundary of the confidence interval.

### 2.4 Computation of the Basic Reproduction Number 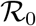

We can also compute the basic reproduction number based on our proposed SuEIR model. Note that our model has a different dynamics from that of SIR and SEIR models. Thus we cannot directly apply the standard computation method of 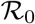 for the SIR or SEIR model to compute such number. Instead, we use the method proposed in Heffernan et al. (2005) to calculate 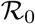 based on the next-generation matrix. In specific, let x = (*x*_1_,…,*x*_4_)^⊤^ with *x_i_* being the number of infected individuals in the compartment *i*. Then we denote by function *F_i_*(x) the rate of new infections in compartment *i*, and denote by 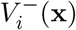 and 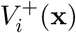 the rate of individuals transferred out of the compartment *i* and the rate of individuals transferred into the compartment *i* by all other means respectively. Let 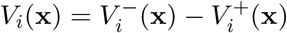, *F*(x) = (*F*_1_(x),…,*F*_4_(x))^⊤^ and *V*(x) = (*V*_1_(x),…,*V*_4_(x))^⊤^ The ODE (1) can be rewritten as *dx/dt* = *F*(*x*) − *V*(*x*) with

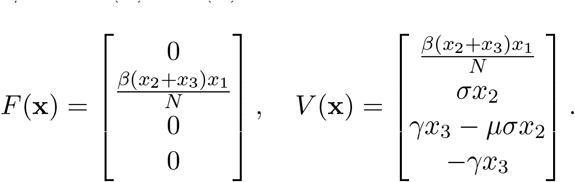

Note that the disease-free equilibrium of our model is x* = (*N*, 0, 0, 0)^⊤^. Let F and V be the partial Jacobian matrices of functions *F*(x) and *V* (x) with respect to the number of individuals in the “infective” compartments (both *E* and *I* compartments in the SuEIR model), i.e., *x*_2_ and *x*_3_, i.e.,

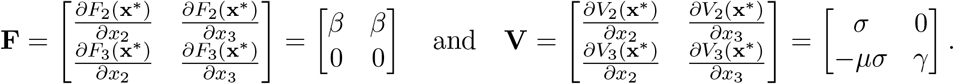

Then the next-generation matrix **G** = **FV**^−1^ can be computed as follows:

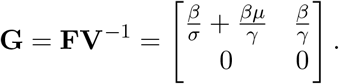

Note that 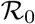 is given by the largest eigenvalue of next generation matrix **G** (Heffernan et al., 2005). Therefore, it is easy to show that the basic reproduction number of our proposed SuEIR model is

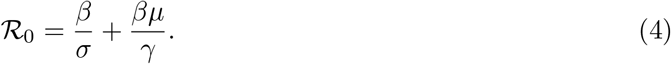

In contrast, the basic reproduction number for SIR and SEIR is 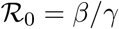.

## 3 Results

In this section, we present the forecast results, including confirmed cases and deaths, peak date and reproduction number, by our method.

Data collection. We use the data from the Johns Hopkins University Center for Systems Science and Engineering^2^ (Dong et al., 2020) to train our model for national-level forecasts. To train the state-level models, we use the data from The New York Times.^3^In addition, we use the data from 03/22/2020 (most states have already issued the stay-at-home order by this date) to 05/10/2020 to train our models. More specifically, we use the reported data from 03/22/2020 to 05/03/2020 to train the SuEIR model while using the data from 05/04/2020 to 05/10/2020 for validation.

Prediction Results. For the interest of space, we present the forecast results of our models for the US and states with more than 40, 000 total cases, including New York, New Jersey, Illinois, Massachusetts, California, Pennsylvania, Michigan, Florida and Maryland. For more forecast results, please refer to our forecast website https://covid19.uclaml.org

Table 1 summarizes the projected death and its corresponding 95% confidence interval in the aforementioned regions from 05/12/2020 to 05/18/2020. The results show that our predictions are very close to the reported data, which suggests that our method performs very well in terms of the death forecasts. We also show the long term death forecasts by our method in Table 2. Our results suggest that by June 30, the projected death for the US is 123.4 × 10^3^ (95% CI 109.7× 10^3^-140.4 × 10^3^).

**Table 1:** Short-term (daily ahead) prediction (×10^3^) of total deaths in the US and states with more than 40,000 total cases. For each region, we present the predicted cumulative fatality cases with a 95% confidence interval. The reported number of deaths (groundtruth) from the JHU CSSE (for the US) and The New York Times (for different states) is presented right below the predictions.

| Date | 05/12 | 05/13 | 05/14 | 05/15 | 05/16 | 05/17 | 05/18 |
| --- | --- | --- | --- | --- | --- | --- | --- |
| US | 82.60<br>[81.75, 83.49] | 84.10<br>[82.82, 85.44] | 85.57<br>[83.86, 87.37] | 87.01<br>[84.88, 89.27] | 88.42<br>[85.87, 91.14] | 89.79<br>[86.83, 92.97] | 91.13<br>[87.76, 94.77] |
|  | 82.38 | 84.12 | 85.90 | 87.53 | 88.75 | 89.56 | 90.35 |
| NY | 27.51<br>[27.27, 27.75] | 27.92<br>[27.57, 28.29] | 28.33<br>[27.86, 28.82] | 28.72<br>[28.14, 29.33] | 29.09<br>[28.41, 29.83] | 29.46<br>[28.67, 30.31] | 29.82<br>[28.92, 30.78] |
|  | 27.28 | 27.44 | 27.61 | 27.75 | 27.95 | 28.16 | 28.30 |
| NJ | 9.568<br>[9.482, 9.656] | 9.719<br>[9.590, 9.852] | 9.866<br>[9.696, 10.04] | 10.00<br>[9.798, 10.22] | 10.14<br>[9.897, 10.41] | 10.28<br>[9.994, 10.58] | 10.41<br>[10.08, 10.76] |
|  | 9.508 | 9.702 | 9.946 | 10.13 | 10.24 | 10.35 | 10.43 |
| IL | 3.592<br>[3.540, 3.648] | 3.685<br>[3.605, 3.770] | 3.776<br>[3.669, 3.893] | 3.867<br>[3.731, 4.015] | 3.956<br>[3.792, 4.137] | 4.044<br>[3.852, 4.258] | 4.130<br>[3.911, 4.378] |
|  | 3.617 | 3.815 | 3.945 | 4.075 | 4.149 | 4.198 | 4.257 |
| MA | 5.151<br>[5.103, 5.199] | 5.234<br>[5.162, 5.308] | 5.315<br>[5.221, 5.414] | 5.395<br>[5.277, 5.518] | 5.472<br>[5.332, 5.620] | 5.548<br>[5.385, 5.720] | 5.621<br>[5.437, 5.817] |
|  | 5.141 | 5.315 | 5.482 | 5.592 | 5.705 | 5.797 | 5.862 |
| CA | 2.846<br>[2.807, 2.887] | 2.913<br>[2.853, 2.981] | 2.982<br>[2.899, 3.079] | 3.052<br>[2.945, 3.181] | 3.124<br>[2.990, 3.287] | 3.196<br>[3.036, 3.397] | 3.270<br>[3.081, 3.510] |
|  | 2.902 | 3.014 | 3.039 | 3.192 | 3.254 | 3.290 | 3.322 |
| PA | 3.917<br>[3.886, 3.948] | 3.970<br>[3.925, 4.017] | 4.022<br>[3.962, 4.084] | 4.073<br>[3.999, 4.149] | 4.122<br>[4.034, 4.212] | 4.169<br>[4.068, 4.273] | 4.215<br>[4.101, 4.333] |
|  | 3.924 | 4.104 | 4.298 | 4.432 | 4.490 | 4.504 | 4.560 |
| MI | 4.705<br>[4.663, 4.747] | 4.776<br>[4.715, 4.840] | 4.846<br>[4.765, 4.930] | 4.914<br>[4.814, 5.018] | 4.980<br>[4.861, 5.104] | 5.044<br>[4.906, 5.187] | 5.105<br>[4.950, 5.268] |
|  | 4.674 | 4.714 | 4.787 | 4.825 | 4.880 | 4.891 | 4.915 |
| FL | 1.782<br>[1.765, 1.799] | 1.811<br>[1.786, 1.838] | 1.840<br>[1.807, 1.876] | 1.869<br>[1.827, 1.913] | 1.897<br>[1.847, 1.950] | 1.924<br>[1.866, 1.986] | 1.951<br>[1.884, 2.022] |
|  | 1.778 | 1.826 | 1.874 | 1.916 | 1.963 | 1.972 | 1.996 |
| MD | 1.727<br>[1.703, 1.751] | 1.768<br>[1.732, 1.805] | 1.808<br>[1.760, 1.860] | 1.848<br>[1.788, 1.914] | 1.888<br>[1.815, 1.968] | 1.926<br>[1.842, 2.021] | 1.965<br>[1.868, 2.073] |
|  | 1.756 | 1.809 | 1.866 | 1.911 | 1.957 | 1.992 | 2.023 |

**Table 2:**
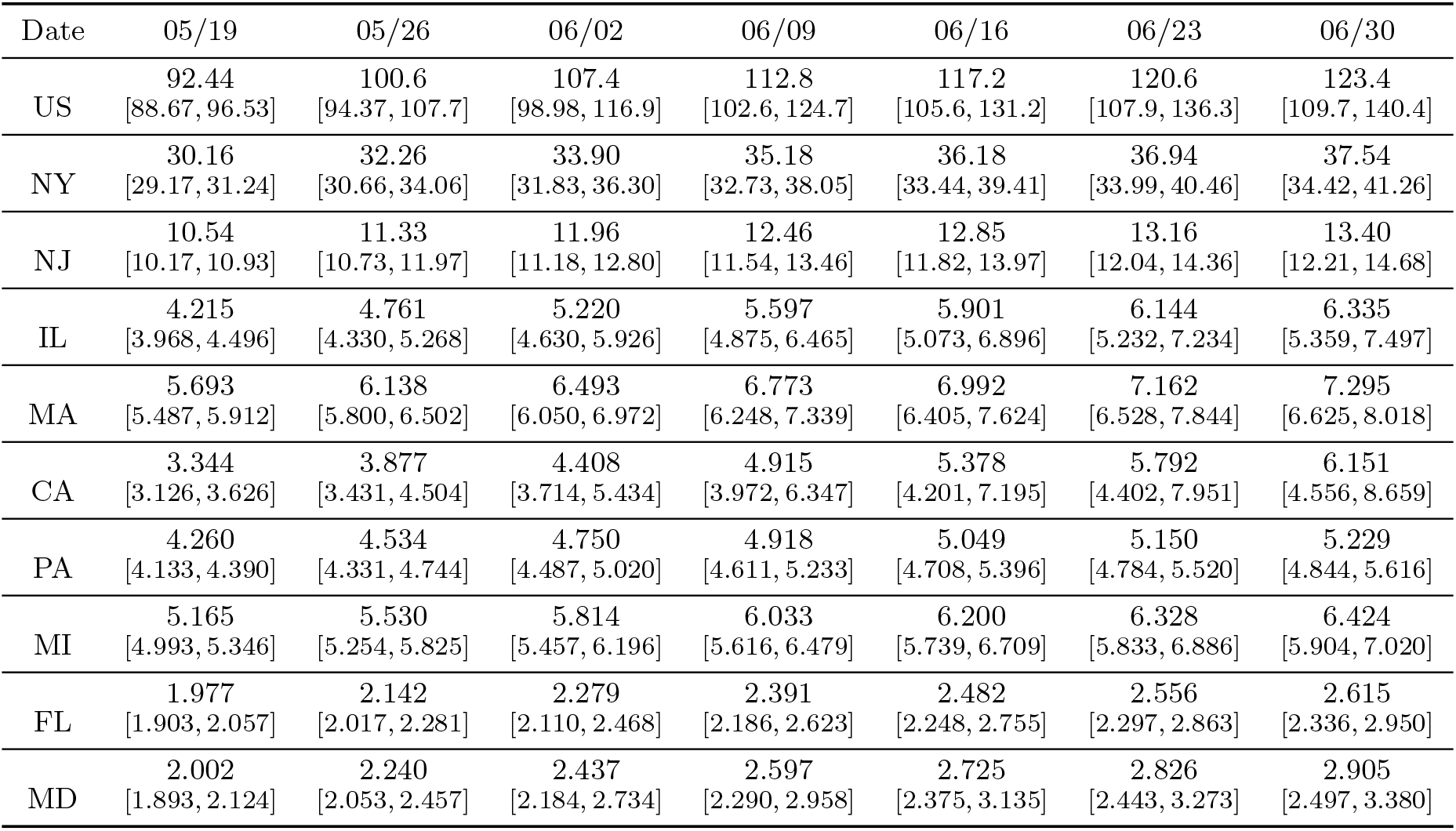
Long-term (weakly ahead) prediction (×10^3^) of total deaths in the US and states with more than 40,000 total cases. For each region, we present the predicted cumulative fatality cases with a 95% confidence interval.

| Date | 05/19 | 05/26 | 06/02 | 06/09 | 06/16 | 06/23 | 06/30 |
| --- | --- | --- | --- | --- | --- | --- | --- |
| US | 92.44<br>[88.67, 96.53] | 100.6<br>[94.37, 107.7] | 107.4<br>[98.98, 116.9] | 112.8<br>[102.6, 124.7] | 117.2<br>[105.6, 131.2] | 120.6<br>[107.9, 136.3] | 123.4<br>[109.7, 140.4] |
| NY | 30.16<br>[29.17, 31.24] | 32.26<br>[30.66, 34.06] | 33.90<br>[31.83, 36.30] | 35.18<br>[32.73, 38.05] | 36.18<br>[33.44, 39.41] | 36.94<br>[33.99, 40.46] | 37.54<br>[34.42, 41.26] |
| NJ | 10.54<br>[10.17, 10.93] | 11.33<br>[10.73, 11.97] | 11.96<br>[11.18, 12.80] | 12.46<br>[11.54, 13.46] | 12.85<br>[11.82, 13.97] | 13.16<br>[12.04, 14.36] | 13.40<br>[12.21, 14.68] |
| IL | 4.215<br>[3.968, 4.496] | 4.761<br>[4.330, 5.268] | 5.220<br>[4.630, 5.926] | 5.597<br>[4.875, 6.465] | 5.901<br>[5.073, 6.896] | 6.144<br>[5.232, 7.234] | 6.335<br>[5.359, 7.497] |
| MA | 5.693<br>[5.487, 5.912] | 6.138<br>[5.800, 6.502] | 6.493<br>[6.050, 6.972] | 6.773<br>[6.248, 7.339] | 6.992<br>[6.405, 7.624] | 7.162<br>[6.528, 7.844] | 7.295<br>[6.625, 8.018] |
| CA | 3.344<br>[3.126, 3.626] | 3.877<br>[3.431, 4.504] | 4.408<br>[3.714, 5.434] | 4.915<br>[3.972, 6.347] | 5.378<br>[4.201, 7.195] | 5.792<br>[4.402, 7.951] | 6.151<br>[4.556, 8.659] |
| PA | 4.260<br>[4.133, 4.390] | 4.534<br>[4.331, 4.744] | 4.750<br>[4.487, 5.020] | 4.918<br>[4.611, 5.233] | 5.049<br>[4.708, 5.396] | 5.150<br>[4.784, 5.520] | 5.229<br>[4.844, 5.616] |
| MI | 5.165<br>[4.993, 5.346] | 5.530<br>[5.254, 5.825] | 5.814<br>[5.457, 6.196] | 6.033<br>[5.616, 6.479] | 6.200<br>[5.739, 6.709] | 6.328<br>[5.833, 6.886] | 6.424<br>[5.904, 7.020] |
| FL | 1.977<br>[1.903, 2.057] | 2.142<br>[2.017, 2.281] | 2.279<br>[2.110, 2.468] | 2.391<br>[2.186, 2.623] | 2.482<br>[2.248, 2.755] | 2.556<br>[2.297, 2.863] | 2.615<br>[2.336, 2.950] |
| MD | 2.002<br>[1.893, 2.124] | 2.240<br>[2.053, 2.457] | 2.437<br>[2.184, 2.734] | 2.597<br>[2.290, 2.958] | 2.725<br>[2.375, 3.135] | 2.826<br>[2.443, 3.273] | 2.905<br>[2.497, 3.380] |

We demonstrate the projected number of confirmed cases by our approach along with its 95% confidence interval in Table 3. The results suggest that our predictions in terms of the number of confirmed cases are also reasonably accurate. For example, the reported number of confirmed cases in the US on 05/18/2020 is 1508 × 10^3^, and our projected number is 1496 × 10^3^ (95% CI 1443 × 10^3^-1572 × 10^3^), which underestimates 12 × 10^3^ cases. In addition, the projected number of long term confirmed case is presented in Table 4. It shows that by 06/30/2020, the projected number of confirmed cases for the US is 1900 × 10^3^ (95% CI 1638 × 10^3^-2362 × 10^3^).

**Table 3:**
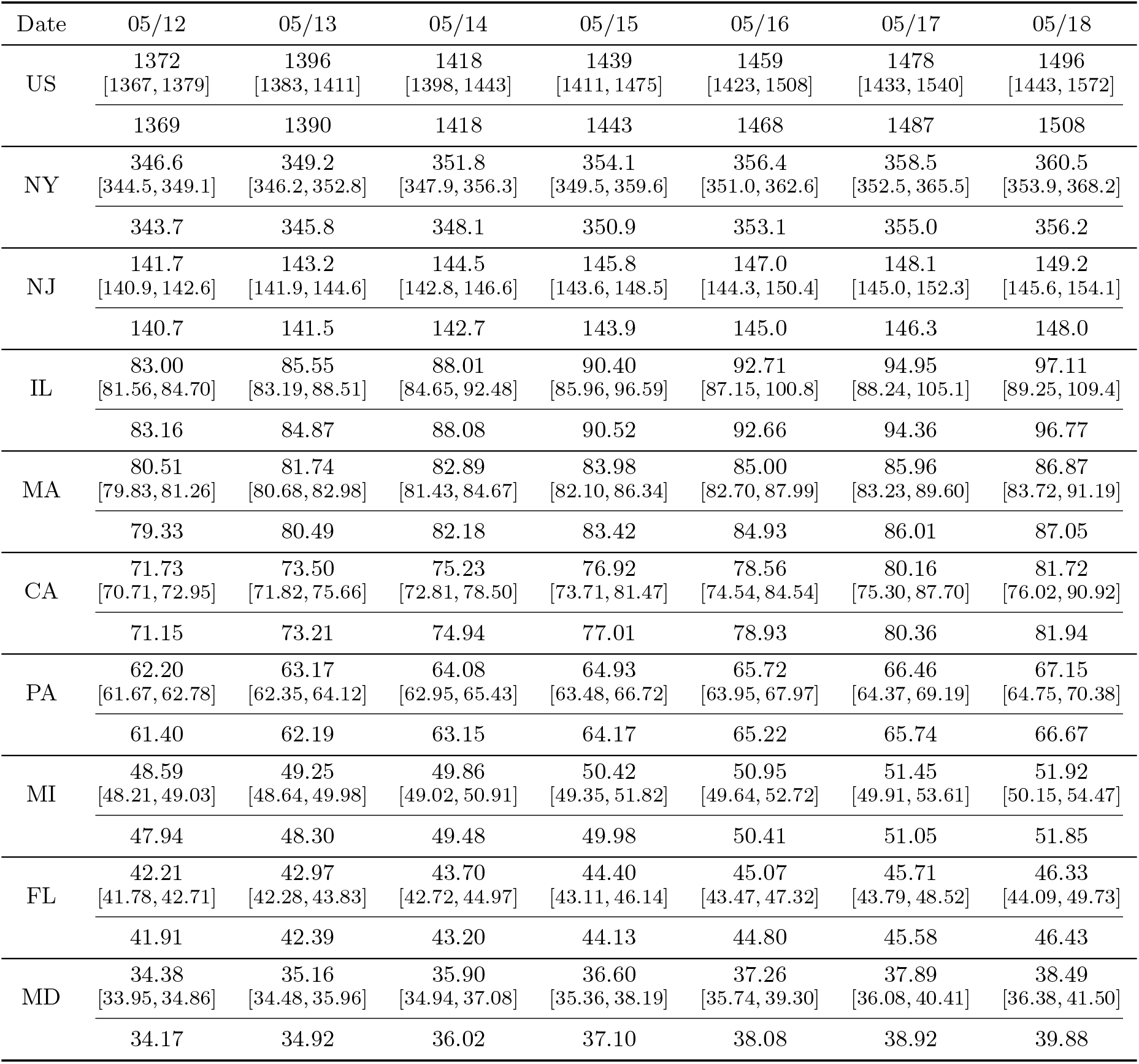
Short-term (daily ahead) prediction (×10^3^) of total confirmed cases in the US and states with more than 40,000 total cases. For each region, we present the predicted cumulative confirmed cases with a 95% confidence interval. The groundtruth number from the JHU CSSE (for the US) and The New York Times (for different states) is presented under the row of predictions.

**Table 4:** Long-term (weakly ahead) prediction (×10^3^) of total confirmed cases in the US and states with more than 40,000 total cases. For each region, we present the predicted cumulative confirmed cases with a 95% confidence interval.

| Date | 05/19 | 05/26 | 06/02 | 06/09 | 06/16 | 06/23 | 06/30 |
| --- | --- | --- | --- | --- | --- | --- | --- |
| US | 1513<br>[1451, 1604] | 1616<br>[1500, 1810] | 1696<br>[1537, 1981] | 1762<br>[1568, 2115] | 1816<br>[1595, 2218] | 1862<br>[1618, 2299] | 1900<br>[1638, 2362] |
| NY | 362.4<br>[355.2, 370.8] | 373.4<br>[362.8, 385.7] | 381.7<br>[368.6, 397.5] | 388.4<br>[372.3, 408.7] | 393.8<br>[375.3, 417.7] | 398.4<br>[377.9, 424.9] | 402.3<br>[380.1, 430.9] |
| NJ | 150.2<br>[146.1, 155.8] | 155.9<br>[148.9, 166.8] | 160.1<br>[150.9, 175.5] | 163.4<br>[152.4, 182.4] | 166.1<br>[153.7, 187.8] | 168.3<br>[154.8, 192.0] | 170.2<br>[155.8, 195.5] |
| IL | 99.20<br>[90.19, 113.8] | 112.1<br>[95.51, 143.7] | 122.6<br>[99.67, 168.8] | 131.2<br>[103.2, 187.9] | 138.4<br>[106.3, 201.8] | 144.5<br>[109.1, 212.1] | 149.6<br>[111.6, 219.8] |
| MA | 87.72<br>[84.17, 92.73] | 92.61<br>[86.51, 102.3] | 96.18<br>[88.14, 109.9] | 98.96<br>[89.43, 115.7] | 101.2<br>[90.52, 120.2] | 103.0<br>[91.46, 123.6] | 104.6<br>[92.28, 126.3] |
| CA | 83.25<br>[76.70, 94.19] | 92.98<br>[80.74, 117.0] | 101.2<br>[84.12, 137.0] | 108.4<br>[87.13, 152.9] | 114.5<br>[89.83, 165.1] | 119.9<br>[92.28, 174.4] | 124.5<br>[94.49, 181.6] |
| PA | 67.80<br>[65.09, 71.53] | 71.43<br>[66.84, 78.67] | 73.99<br>[67.99, 84.21] | 75.96<br>[68.88, 88.47] | 77.56<br>[69.63, 91.77] | 78.90<br>[70.28, 94.36] | 80.05<br>[70.86, 96.44] |
| MI | 52.36<br>[50.37, 55.31] | 54.93<br>[51.58, 60.57] | 56.87<br>[52.45, 64.73] | 58.39<br>[53.14, 67.94] | 59.61<br>[53.70, 70.41] | 60.60<br>[54.16, 72.32] | 61.41<br>[54.54, 73.81] |
| FL | 46.92<br>[44.37, 50.94] | 50.59<br>[45.94, 59.21] | 53.61<br>[47.18, 66.52] | 56.19<br>[48.27, 72.57] | 58.43<br>[49.25, 77.44] | 60.41<br>[50.14, 81.36] | 62.16<br>[50.96, 84.54] |
| MD | 39.06<br>[36.66, 42.58] | 42.31<br>[38.11, 49.50] | 44.68<br>[39.07, 54.95] | 46.50<br>[39.83, 59.08] | 47.96<br>[40.47, 62.16] | 49.17<br>[41.03, 64.47] | 50.20<br>[41.51, 66.26] |

Our method can also forecast the peak date in different regions, i.e., the date with the largest number of active cases, as shown in Table 5. It can be seen that the projected peak date of the US is 06/01/2020, New York state is 05/10/2020, New Jersey is 05/19/2020, Illinois is 06/07/2020, Massachusetts is 05/23/2020, California is 07/01/2020, Pennsylvania is 05/20/2020, Michigan is 05/11/2020, Florida is 06/14/2020, Maryland is 05/27/2020.

**Table 5:**
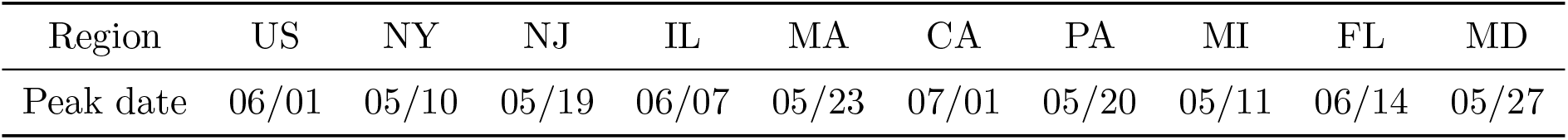
Projected peak date by our model in different regions.

Table 6 summarizes the basic reproduction number 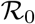 estimated by (4) in different regions, which characterizes the spread of the virus at the beginning of the epidemic. The results vary for different states, which are consistent with the severity of the coronavirus outbreak in these regions since mid March. For example the 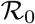 values of the states in the Northeastern US (e.g., NY: 3.6, NJ: 4.5, MA:4.2) are significantly higher than those of other states (e.g. CA: 2.2, MI: 2.1, FL: 2.4).

**Table 6:** Estimated basic reproduction number 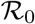 by our model in different regions.

| Region | US | NY | NJ | IL | MA | CA | PA | MI | FL | MD |
| --- | --- | --- | --- | --- | --- | --- | --- | --- | --- | --- |
| $\mathcal{R}_0$ | 2.5 | 3.6 | 4.5 | 3.6 | 4.2 | 2.2 | 3.3 | 2.1 | 2.4 | 2.9 |

## 4 Discussion

We developed a novel epidemic model called SuEIR to infer the unreported cases of individuals contacting COVID-19. Based on this new model, we further develop a machine learning approach to forecast the numbers of confirmed and fatality cases in the US.

Our model can provide accurate short-term (daily ahead) projections for both confirmed cases and fatality cases at national and state levels, which demonstrates its effectiveness. In the long term, the prediction results by our model suggest that the numbers of confirmed cases and death will keep increasing rapidly within one month. In particular, at the end of June, our model forewarns that there will be approximately 2 millions confirmed infectious cases and 120K reported deaths in the United States.

Our model uses training data since 03/22/2020 at which most states have already issued stay-at-home order, and assumes that the contact rate maintains the same level during the training and prediction period. However, starting in May, many states have already lifted the restrictions of businesses and public spaces and considered reopening that allows people to go back to restaurants and offices and places of worship. It remains unclear how these reopening orders affect the contact rate as well as the spread of the virus and therefore our current model does not take this into consideration.

Moreover, we found that for most states, the learned discover rate (i.e., *μ*) is less than 0.1, which implies that a large fraction of “Exposed” individuals will finally recover/die without being tested and reported. This further suggests that the actual number of infected cases in the US may be more than 10 millions, while most of them are not counted. This result is consistent with the recent findings by the researchers from the University of Southern California (Sood et al., 2020), which show that 4.65% (CI: [2.8%, 5.6%]) of Los Angeles residents have already contracted the COVID-19 virus, which is approximately 23 times more than the official reported numbers.

## Data Availability

All data are publicly available.

https://github.com/CSSEGISandData/COVID-19

https://github.com/nytimes/covid-19-data

## Footnotes

1 Here we omit the numbers of removed cases and recovered cases at the initialization by setting 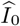 and 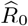 to be the reported numbers of confirmed cases and fatality cases.

2 https://github.com/CSSEGISandData/COVID-19

3 https://github.com/nytimes/covid-19-data

